# Automated EEG Classification to Track Levels of Consciousness

**DOI:** 10.64898/2026.06.18.26355981

**Authors:** William H. Curley, Andrew Hoopes, David W. Zhou, Mary M. Conte, Jonathan D. Victor, Nicholas D. Schiff, Brian L. Edlow

## Abstract

Precise prognostication in acute brain injury is limited by a lack of reliable biomarkers of consciousness available to clinicians at the bedside. The ABCD framework is a method of classifying resting-state clinical EEG into categories that reflect levels of thalamocortical network function. ABCD classifications in the intensive care unit (ICU) have been shown to provide diagnostic and prognostic utility for patients with severe brain injuries, but the current gold standard for ABCD classification is visual inspection of power spectra, which is labor-intensive and requires expertise in spectral analysis. Using 4,611 manually classified EEG power spectra, we developed an automated, highly accurate, and well-calibrated convolutional neural net-based classifier of EEG into ABCD categories. The classifier has performance comparable to that of the current gold standard and that outperforms an alternative method of automated spectral analysis. As proof-of-principle for clinical implementation, we apply the classifier to a continuous EEG record from a patient with acute severe traumatic brain injury in the ICU, demonstrating its ability to yield continuous ABCD classifications that capture state fluctuations with high temporal and spatial resolution. The automated ABCD classifier allows for efficient analysis of continuous EEG records, facilitating the translation of the ABCD framework to the bedside for patients with acute severe brain injuries. The ABCD classifier also creates new opportunities to efficiently analyze large EEG datasets and generate new insights into the electrophysiological properties of human consciousness.

## Introduction

Diagnosis and prognosis for patients with severe brain injuries is challenging, particularly in the acute phase of injury. Accurate assessment of acutely brain-injured patients in the intensive care unit (ICU) is complicated by multiple factors including a high rate of misdiagnosis when using behavioral measures alone, the potential for cognitive motor dissociation (CMD), rapid fluctuations in arousal and awareness, sedation, and comorbid critical illness (Schnakers et al., 2009; Schiff, 2015; Edlow et al., 2021). Accordingly, there is a need for reliable and reproducible tools to support evaluation of patients with acute brain injuries in the ICU. Approaches that leverage routinely acquired data, such as resting-state electroencephalography (EEG), are particularly well suited for bedside integration and real-time clinical use (Vespa et al., 2002; Hebb et al., 2007; Forgacs et al., 2017; Tolonen et al., 2018; Alkhachroum et al., 2020; Forgacs et al., 2020, 2022; Amiri et al., 2023; Curley et al., 2022a,b; Zhou et al., 2024).

One approach to EEG-based assessment in acute brain injury is classifying EEG power spectral estimates according to the ABCD framework (Forgacs et al., 2017, 2020, 2022; Curley et al., 2022a). Derived from the mesocircuit model of disorders of consciousness, this framework organizes EEG power spectral estimates into four categories that correspond to progressive, widely separated levels of thalamocortical network function, as demonstrated in Figure 1 (Schiff, 2010, 2016).

**Figure 1.**
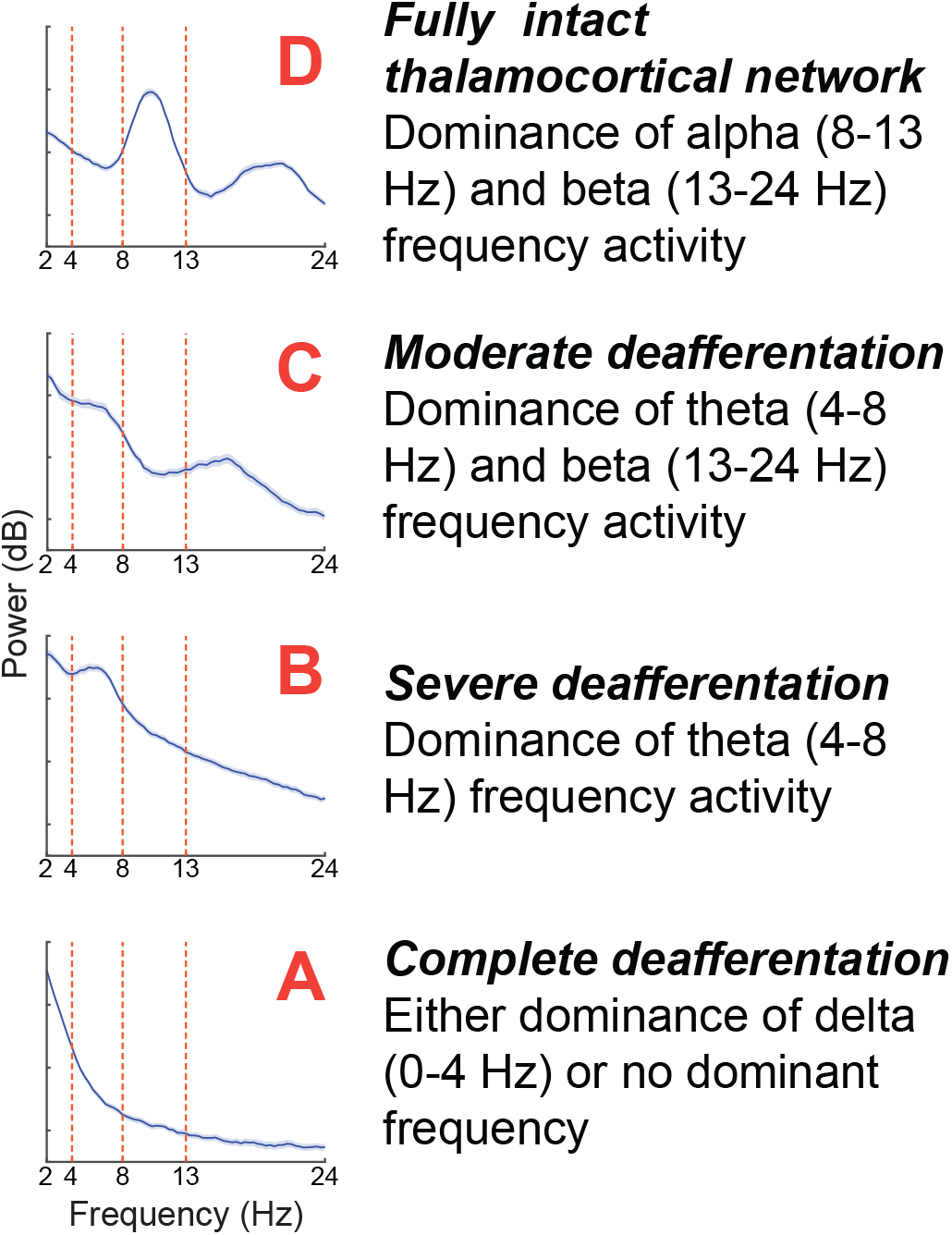
Relationship between ABCD framework categories and thalamocortical network integrity. *D*-type spectra correspond to a fully intact and functional thalamocortical network while *C, B*, and *A*-type spectra represent stepped down levels of thalamocortical network functioning in descending order.

The ABCD categories and their physiologic correlates according to the mesocircuit model are as follows (Curley et al., 2022a). The *A* category (no spectral peak above the delta [<4 Hz] range) corresponds to complete disconnection of the thalamocortical network wherein neocortical neurons are hyperpolarized and produce only low frequency oscillations, as in the “cortical slab preparation” (Timofeev et al., 2000). In the *B* category (spectral peak in the theta [4-8 Hz] range), the thalamocortical network is severely but not completely disconnected, resulting in comparatively less polarization of neocortical neurons and allowing for theta frequency bursting (Silva et al., 1991). In the *C* category (spectral peaks in theta and beta [13-24 Hz] range), connections between thalamus and cortex are only moderately disconnected and thus, thalamic neuronal bursting drives simultaneous theta and beta oscillations in connected cortex (Llinás et al., 1999, 2005). The *D* category (spectral peaks in the alpha [8-13 Hz] and beta ranges) represents a normal, completely intact thalamocortical network wherein normal thalamic and cortical activity produces alpha and beta oscillations as can be seen in the spectral profile of a healthy, awake adult EEG (Schomer and Lopes da Silva, 2017).

The thalamocortical network is integral to supporting consciousness (Redinbaugh et al., 2020; Janson et al., 2021), and its disruption has been implicated in the patho-physiology of disorders of consciousness across diverse etiologies (Fridman et al., 2014; Lutkenhoff et al., 2015; Fischer et al., 2016; Coulborn et al., 2021). Consistent with this model, ABCD classification of EEG correlates with outcome in acute subarachnoid hemorrhage (Forgacs et al., 2022) and anoxic brain injury following cardiac arrest (Forgacs et al., 2017). Application of the ABCD framework is also useful in the evaluation of patients in the acute and chronic phases of traumatic brain injury (TBI), specifically with regard to diagnosis and capturing rapid fluctuations in brain function that are otherwise imperceptible through bedside examinations (Curley et al., 2022a). As such, translation of real-time ABCD classification in the ICU has potential to improve the detection of patients with CMD through guiding assessment timing (i.e. behavioral assessments, task-based EEG assessments) based on when patients are in their optimal state to demonstrate signs of consciousness.

However, current applications of the ABCD framework rely on manual classification of EEG spectra, which is time-intensive, subjective, and requires expert interpretation. Most critically, this process is performed offline,limiting its use for informing on-demand clinical assessment.

To address this gap, we developed a convolutional neural network (CNN) classifier that rapidly assigns per-channel ABCD categories from EEG spectral estimates. The model achieves performance comparable to expert visual classification, despite known inter-rater variability, and provides well-calibrated probability estimates that reflect prediction accuracy. Our approach substantially outperforms a commonly used peak-fitting–based method for spectral analysis. Finally, we demonstrate its application to continuous EEG data from an acutely brain-injured patient as a proof-of-concept for clinical deployment, enabling high-temporal-resolution tracking of state fluctuations. This automated approach supports scalable, reproducible analysis and advances the translation of the ABCD framework toward routine clinical use to improve care for patients with acute brain injuries.

## Materials and Methods

### Datasets

The datasets used to develop and evaluate our classification framework were obtained from two previously conducted and published studies. The *internal* cohort (Edlow et al., 2017; Curley et al., 2022a) includes EEG recordings from 16 healthy controls and 20 patients admitted to the Massachusets General Hospital intensive care unit (ICU) with acute traumatic coma. The second, *external* cohort from Weill Cornell Medicine (Bodien et al., 2024) includes EEG recordings from 28 patients enrolled in a longitudinal study of recovery following severe, non-progressive brain injury. The external cohort was used exclusively as a held-out evaluation dataset and not used for model development.

Participants contributed at least one 5-minute segment of resting-state EEG acquired at sampling frequencies of 200, 250, or 256 Hz. Recordings involved either 19 electrodes positioned according to the international 10–20 system (internal cohort) or 37 electrodes arranged according to an augmented 10–20 system with 18 additional electrodes (external cohort).

### Spectral Processing

Continuous EEG data were detrended, bandpass filtered between 1 and 30 Hz using a zero-phase, third-order Butterworth filter, segmented into 3-second epochs, and rereferenced to a Hjorth Laplacian montage to enhance spatial localization of cortical signals (Hjorth, 1975; Thickbroom et al., 1984). Power spectral density (PSD) estimates were computed on a channel-by-channel basis for each epoch using a multi-taper spectral estimation approach (Thomson, 1982; Percival and Walden, 1993; Bokil et al., 2010). Five tapers were used, yielding a spectral resolution of 2 Hz with estimates spaced at 1/3 Hz intervals. Spectra were averaged across epochs within each resting-state block and analyzed over the 2–24 Hz frequency range, consistent with prior applications of the ABCD framework (Forgacs et al., 2017).

Epochs contaminated by electromyographic activity, eye-blink artifact, or electrical interference were excluded from analysis, and entire resting-state blocks were discarded when artifact burden precluded reliable spectral estimation. This artifact rejection was performed manually by a single investigator (WC) (Forgacs et al., 2014, 2017; Curley et al., 2018). An average of 40.2% of epochs were excluded per block for healthy controls and 60.9% for patients. In total, we derived 4,472 and 139 individual channel-level resting-state power spectra from the internal and external cohorts, respectively.

### Manual Labeling

We manually assigned each power spectrum from all available EEG channels to one of the ABCD categories: *A, B, C*, or *D*, according to the framework previously described and illustrated in Figure 1 (Schiff, 2016) and previously published (Curley et al., 2022a). All spectra not conforming to the minimum requirements of any one category (e.g. an alpha peak without an accompanying beta peak) or demonstrated evidence of artifact, including electrical interference, such as square-wave patterns, or electromyogenic contamination, were classified as ‘other’ or *Z*. Prior to labeling, all spectra were anonymized, randomly shuffled, and stripped of clinical variables.

As previously reported, visual inspection-based classification has been demonstrated to have substantial interrater (89% concordance, Fleiss’s *κ* = 0.64) and intra-rater (Fleiss’s *κ* = 0.70) agreement (Meys et al., 2025; Forgacs et al., 2017). As such, internal cohort spectra labeling was performed by a single investigator (WC) who was blinded to all clinical variables, did not have any patient contact, and was not involved in data collection. This investigator also noted whether peaks were present within each of the pre-defined frequency bands (delta, theta, alpha, beta), for each spectrum. Four expert raters independently labeled spectra from the external cohort (DZ, MC, JV, NS), each assigning a single label per spectrum and all of whom were blinded to all clinical variables. We selected a spectrum’s ground truth label as the most frequently chosen label across raters. Spectra without a majority label (i.e., no label assigned by at least three of four raters) were assigned a consensus label following group discussion. We quantified inter-rater reliability using Fleiss’s *κ*.

Spectra from 13 controls and 15 patients in the internal cohort were reserved for model training, and the remaining spectra from both cohorts were held out for evaluation. The distribution of labeled spectra across datasets and label categories is detailed in Table 1 (bottom).

**Table 1.** **Top:** Per-category inter-rater agreement across four raters of the external cohort, reported as exact (unanimous) and majority agreement. Categories are based on the final groundtruth label, assigned by majority vote or consensus discussion when no majority existed. **Bottom:** Per-category distribution of channel-level power spectra used separately for model training, validation, and evaluation.

| Agreement | A | B | C | D | Z |
| --- | --- | --- | --- | --- | --- |
| Exact | 77.2% | 50.0% | 0.0% | 28.6% | 58.6% |
| Majority | 94.7% | 79.4% | 50.0% | 85.7% | 86.5% |

**Table 1. Top:** Per-category inter-rater agreement across four raters of the external cohort, reported as exact (unanimous) and majority agreement.
| Data Split | A | B | C | D | Z |
| --- | --- | --- | --- | --- | --- |
| Internal Train | 155 | 237 | 431 | 1,554 | 800 |
| Internal Validation | 20 | 28 | 50 | 200 | 80 |
| Internal Evaluation | 66 | 53 | 149 | 477 | 172 |
| External Evaluation | 56 | 36 | 6 | 14 | 27 |

### Convolutional Neural Network Classifier

We developed a convolutional neural network (CNN) that takes a single-channel EEG power spectrum as input and outputs a probabilistic classification across five categories: *A, B, C, D*, and the non-conforming class *Z*.

The multiscale convolutional network comprises five sequential residual blocks. Each block includes four 1D convolutional layers operating along the frequency axis, with kernel size 3 and 32 feature channels. Within each block, intermediate convolutional outputs are activated using leaky rectified linear units (Leaky ReLU), and a residual (skip) connection adds the block input to the output of the final convolution. Each residual block is followed by 2*×* max pooling along the frequency axis, resulting in progressive spectral downsampling. The output of the final convolutional block is flattened and passed to a multilayer perceptron consisting of a fully connected layer with 128 units and Leaky ReLU activation, followed by a final linear layer with five output units corresponding to each category. The resulting logits are converted to class probabilities using a softmax operation. Network capacity was manually selected from a coarse architecture search space to maximize validation accuracy.

Prior to input into the network, each power spectrum was cropped to the 2–24 Hz frequency range, resampled to a uniform frequency spacing of 0.23 Hz, and independently min–max normalized to the range [0, 1]. The model was implemented in PyTorch (Paszke et al., 2019) and trained using the Adam optimizer (Kingma and Ba, 2014) with a 10^*−*3^ learning rate. Training was performed with a batch size of 256, using uniform sampling across label categories to address class imbalance. At each optimization step, model parameters were updated by minimizing the categorical cross-entropy loss between predicted class probabilities and ground-truth labels. Optimization was carried out for 10^3^ steps, and model parameters achieving the highest validation accuracy were used for evaluation.

### Simulating Training Spectra

We implemented a mechanism to generate synthetic power spectra *during training* to complement the limited number of manually labeled spectra. This approach was designed to expose the model to a broader range of plausible spectral patterns and thereby improve robustness to diverse, real data encountered *at inference* (Tobin et al., 2017; Tremblay et al., 2018).

Synthetic spectra were generated using a parametric, inverse-modeling approach based on established representations of EEG power spectra as the linear combination of an aperiodic background component and superimposed oscillatory peaks (Donoghue et al., 2020; He et al., 2010). In this framework, the aperiodic component captures the broadband structure of EEG spectra, and oscillatory activity is modeled as Gaussian-shaped peaks centered within canonical frequency bands.

To generate a synthetic spectrum corresponding to a given category label, we first sampled an aperiodic background component modeled as a power-law decay of spectral power with increasing frequency, parameterized by a randomly sampled offset and exponent. Next, we added one or more Gaussian-shaped oscillatory peaks at frequencies characteristic of the target label (e.g., delta, theta, alpha, or beta bands). For each peak, center frequency, bandwidth, and amplitude were independently sampled from predefined, band-specific ranges. Finally, we added zero-mean Gaussian noise with smooth correlations across neighboring frequency bins to introduce additional variability.

During training, simulated spectra were mixed with real EEG spectra and comprised 50% of each training batch. Further details of the simulation model, optimal parameter sampling ranges, and an analysis evaluating the impact of this simulation strategy on performance are provided in the Supplementary Materials.

### FOOOF Baseline

As a performance baseline, we implemented a rule-based classifier via the widely used FOOOF (Fitting Oscillations and One-Over-F) framework for parameterizing EEG power spectra (Donoghue et al., 2020). FOOOF was applied to each channel-level power spectrum to identify oscillatory peaks, which were then mapped to categories within the ABCD framework based on their presence in relevant frequency bands.

FOOOF models each power spectrum by separating the broadband aperiodic component from superimposed oscillatory peaks, with peak detection governed by a set of user-defined fitting hyperparameters. These include the maximum number of allowable peaks, minimum and absolute peak height thresholds, peak width limits, aperiodic mode, and the frequency range over which fitting is performed.

To optimize baseline performance for ABCD classification, we conducted multidimensional grid searches over FOOOF hyperparameters. We evaluated two approaches, described as follows.

In the first approach, we optimized hyperparameters directly for classification performance. For each parameter combination, FOOOF-derived peak detections were mapped to ABCD and non-conforming (Z) categories, and performance was quantified using mean class-balanced recall across the five label categories. The parameter set yielding the highest mean class-balanced recall on the training data was selected.

In the second approach, we optimized hyperparameters separately for peak detection within each frequency band (delta, theta, alpha, beta). Using manually annotated peak presence labels from the training dataset, we identified the parameter combination that maximized peak detection accuracy within each band. The resulting band-specific peak detections were then combined to derive ABCDZ classifications.

The first approach yielded superior classification performance on the training spectra and was used for all baseline evaluations. The full hyperparameter search space and optimal hyperparameters are further detailed in the Supplementary Materials.

## Results

### Inter-rater Reliability of Visual Classification

We quantified inter-rater reliability (Fleiss’s *κ* = 0.62) and found exact agreement among all raters for 56.1% of spec-tra and majority agreement (at least three of four raters) for 82.7%. Per-category agreement rates differ substantially and are summarized in Table 1 (top).

### Classification Performance

We evaluated classification performance on held-out data from both the internal cohort and the independent external cohort, with the latter providing a test of generalization across institution, acquisition protocol, and labeling methodology. As shown in Figure 2A and summarized in Table 2, our method achieved consistent performance across cohorts, with mean class-balanced recall (true positive) of 0.77 on both internal and external evaluation data and comparable macro F1. Inter-rater agreement in the external cohort (Fleiss’s *κ* = 0.62) reflects appreciable variability in expert visual labeling and provides context for interpreting model performance (Table 1).

**Table 2.** **Top:** Per-class classification performance for our CNN method pooled across both evaluation cohorts. Precision denotes positive predictive value; support denotes class sample size. **Bottom:** Mean (class-balanced) recall and macro F1 comparing our CNN method to the FOOOF baseline for each cohort.

| Class | Precision | Recall | F1 | Support |
| --- | --- | --- | --- | --- |
| A | 0.71 | 0.81 | 0.76 | 123 |
| B | 0.66 | 0.76 | 0.71 | 87 |
| C | 0.69 | 0.75 | 0.72 | 155 |
| D | 0.95 | 0.80 | 0.87 | 491 |
| Z | 0.59 | 0.70 | 0.65 | 200 |

**Table 2. Top:** Per-class classification performance for our CNN method pooled across both evaluation cohorts.
| Method | Internal Cohort |  | External Cohort |  |
| --- | --- | --- | --- | --- |
|  | Recall | Macro F1 | Recall | Macro F1 |
| CNN | 0.77 | 0.72 | 0.77 | 0.70 |
| FOOOF | 0.43 | 0.39 | 0.49 | 0.47 |

**Figure 2.**
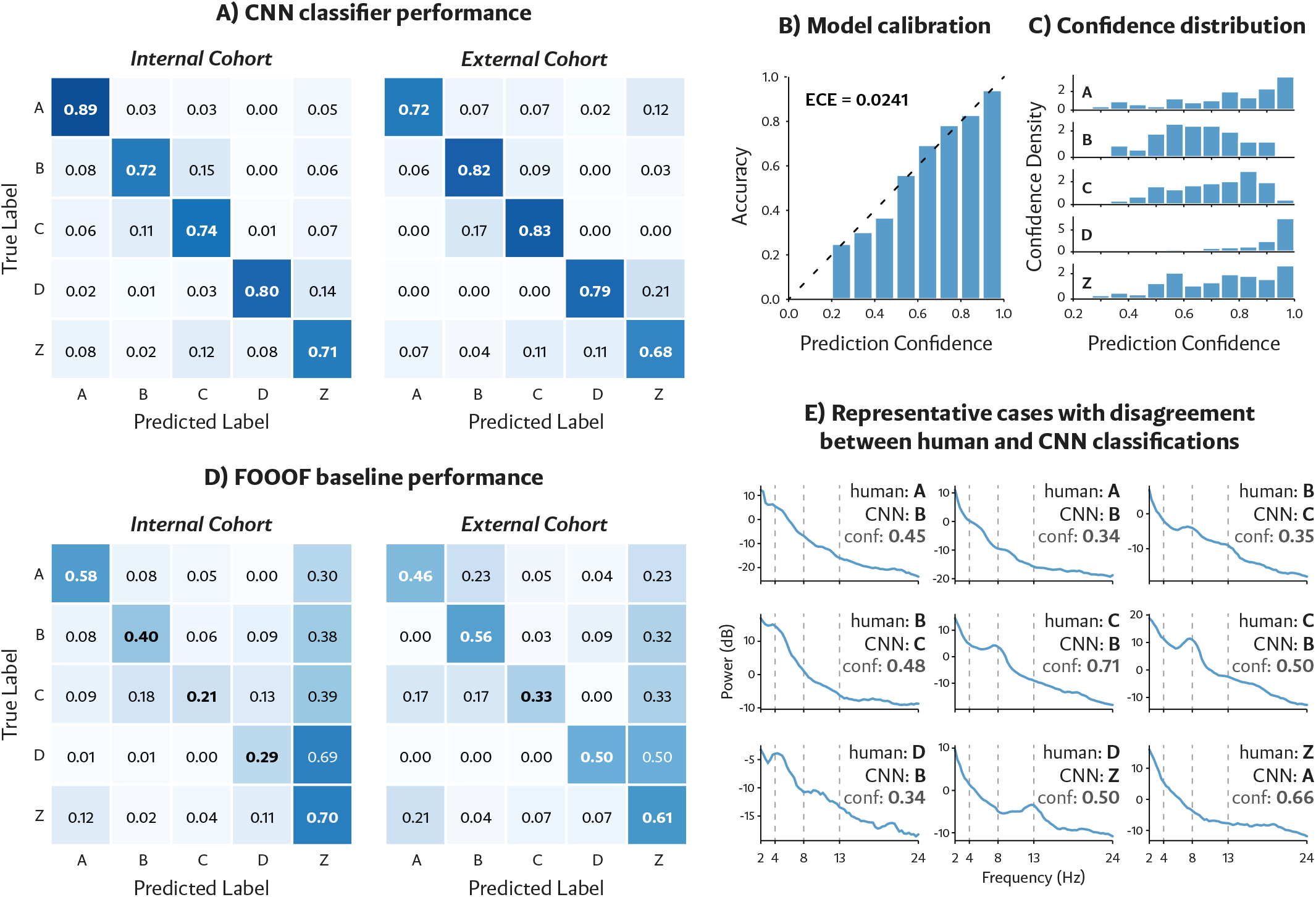
Performance of automated CNN-based ABCD classification. **A:** Confusion matrices demonstrating high classification accuracy of our CNN classifier on the internal (left) and external (right) evaluation cohorts. **B:** Model prediction calibration curve on the evaluation cohort, showing close agreement between predicted confidence and empirical accuracy. **C:** Mean prediction confidence by label category, with lower confidence for B and C categories reflecting increased ambiguity between these adjacent spectral categories. **D:** Confusion matrices for the FOOOF baseline, showing substantially reduced *A, B, C*, and *D* classification accuracy relative to our approach. **E:** Representative cases with disagreement between human and automated CNN classifications including CNN confidence (conf). Disagreements often occurred when spectra exhibited features that were borderline between categories, were not clearly peaks, or did not conform to the ABCD framework.

We additionally evaluated the correspondence between model-predicted probabilities, which can be interpreted as prediction confidence, and empirical accuracy. As shown in Figure 2B, calibration error was low (ECE = 0.0241), indicating close agreement between predicted confidence and observed correctness. Prediction confidence varied systematically across categories (Figure 2C), with lower confidence for B and C categories, which also exhibited lower exact and majority inter-rater agreement.

Representative classification errors are illustrated in Figure 2E. Errors occurred more frequently for spectra with lower inter-rater agreement: mean agreement among raters was higher for correctly classified spectra than for misclassified spectra (86.8% vs. 78.6%). Disagreement among human raters often occurred when spectra were borderline between two categories, contained features that disrupted the spectral baseline but were not clearly peaks, or contained features that did not conform to the spectral patterns delineated in the ABCD framework. When restricting evaluation to external cohort spectra with majority inter-rater agreement, mean class-balanced recall was similar (0.78 versus 0.77). However, restricting further to spectra with exact agreement yielded an increased mean recall of 0.84.

### Comparison to FOOOF Baseline

We compared our classifier to a rule-based baseline using FOOOF-derived spectral features, optimized as described in the Methods. As shown in Figure 2D and Table 2, our CNN method substantially outperformed the FOOOF baseline on both cohorts, with an average absolute accuracy improvement of 30.7 percentage points across evaluation datasets. FOOOF errors were characterized by a strong bias toward the non-conforming (*Z*) category, resulting in poor discrimination among the A–D classes. Across evaluation data, recall for A–D under FOOOF was less than half that achieved by our method on average, whereas performance for Z was comparatively preserved (Table 2).

### Use Case: Continuous EEG Classification

In order to demonstrate the potential for translation of the ABCD framework to the bedside our automated classifier provides, we applied it to over 10 hours of continuous EEG collected from a patient with acute severe TBI in the ICU (Figure 3). We deployed our CNN classifier to assign channel-level ABCD classifications every 5 minutes. As previously published, we then coded each ABCD classification numerically (A=1, B=2, C=3, D=4) and computed a cross-channel average to generate an ABCD index at each timepoint (Curley et al., 2022a). Middle and bottom panels of Figure 3 demonstrate sections of qualitatively increased temporal variability with accompanying topo-graphic heatmaps at representative timepoints demonstrating channel-level ABCD classifications. Overall, we were able to resolve rapid and drastic state fluctuations, at some points occurring over the span of minutes and with spatial heterogeneity.

**Figure 3.**
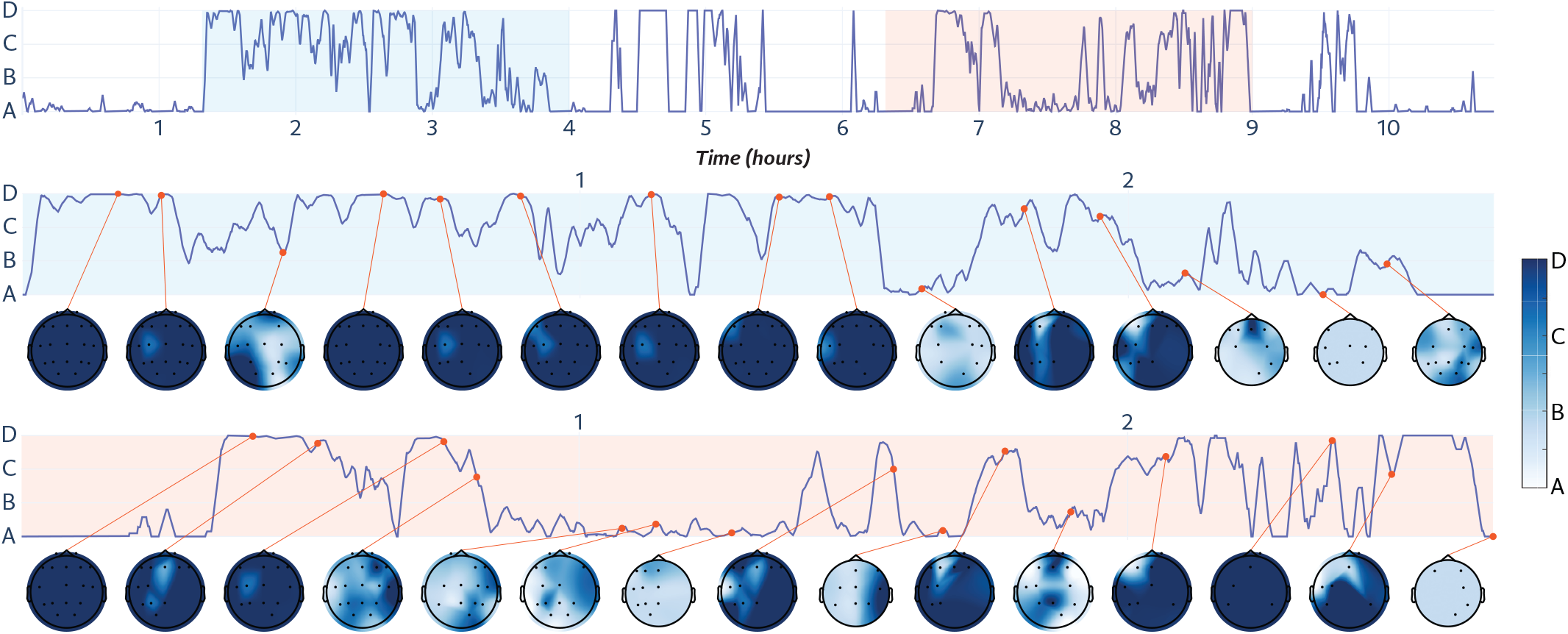
Application of automated CNN classifier to 10+ hours of continuous EEG in a patient with acute severe TBI in the ICU. Top panel demonstrates the ABCD index, an average across channel-level ABCD classifications, at 5-minute intervals. Middle and bottom panels contain callouts corresponding to different sections of the longer continuous timespan in the top panel (indicated by blue and orange highlighting), along with topographic heatmaps of channel-level ABCD classifications at representative timepoints (with colorbar on right denoting colors corresponding to ABCD categories). Included electrodes for each heatmap are represented by black dots, missing channels were classified as ‘Z,’ either because of artifact corruption or spectral features not corresponding to any of the ABCD categories.

## Discussion

We developed an automated approach for classifying EEG spectra according to the ABCD framework, motivated by the limitations of manual interpretation. The model achieved performance comparable to expert visual classification, t he c urrent g old s tandard f or A BCD classification, and outperformed a commonly used automated spectral analysis platform (FOOOF) (Donoghue et al., 2020). We demonstrated its application to continuous EEG, enabling tracking of state fluctuations over time. This approach improves efficiency without compromising accuracy, supports large-scale analyses, and facilitates clinical implementation of the ABCD framework, a measure that already has demonstrated diagnostic and prognostic utility in multiple etiologies of severe brain injury (Forgacs et al., 2017, 2022; Curley et al., 2022a).

The ABCD framework has already been demonstrated to have prognostic utility in hypoxic-ischemic injury and subarachnoid hemorrhage (Forgacs et al., 2017, 2022). The framework additionally has demonstrated diagnostic utility in acute severe TBI (Curley et al., 2022a) but its prognostic utility in this population and other etiologies of severe brain injury has yet to be borne out. Our classifier will facilitate the efficient analysis of large-scale datasets in these additional etiologies of severe brain injury, allowing for assessment of the prognostic utility of the ABCD framework in these populations.

Our classifier allows for analysis of prolonged, continuous EEG records (Figure 3). This could be particularly useful in the clinical evaluation of patients with acute brain injuries, who are known to exhibit marked fluctuations in state (Schnakers et al., 2009; Casali et al., 2013; Gibson et al., 2014; Piarulli et al., 2016; Wannez et al., 2017; Giacino et al., 2018; Demertzi et al., 2019; Kondziella et al., 2020; Curley et al., 2022a). The presence of state fluctuations is likely a contributing factor to the high false negative rate (6/9) for stimulus-based, active EEG tests (i.e., motor imagery tasks) observed in this patient popula-tion (Edlow et al., 2017), especially since successful motor imagery task performance can be state-dependent (Curley et al., 2018). Given that the presence of covert command following in the acute period of injury is associated with better prognosis (Claassen et al., 2019), improved sensitivity for these tests is crucial. By integrating continuous, automated ABCD classification at the bedside of patients with severe brain injuries, clinicians could be guided as to the optimal timing of assessments to improve diagnostic accuracy, improve efficiency of attaining accurate diagnoses and prognoses, and potentially increase the sensitivity of assessments.

We achieved automated classifier accuracy comparable to that of inter-rater reliability of human classifications, the current gold standard for assigning ABCD classifications. Examining cases of human versus automated classifier disagreement highlights the inherent difficulty of assigning ABCD classifications in some cases where the ground truth is not clear (Figure 2E). Disagreement among human raters often occurred when spectra were borderline between two categories or contained features that disrupted the spectral baseline but were not clearly peaks. Additionally, patients with acute severe TBI demonstrate spectral patterns that do not conform to the ABCD framework 25% of the time (Curley et al., 2022a), although all patients in this study exhibited conforming spectral patterns in at least some EEG channels. The significance of non-conforming patterns is not yet clear and further study is required to better understand if addi-tional patterns have clinical relevance. Regardless, when channel-level spectra cannot be accurately categorized, it is likely best for them to be excluded from analysis. Our classifier is well calibrated, which means that prediction confidence is highly associated with accuracy. Thus, applying a confidence threshold for assigning classifications would allow for exclusion of channels unable to be classified with a reasonable degree of confidence. As an example, applying a confidence threshold of 0.5 would prevent truly equivocal cases from being included in analyses while preserving the majority of data (Figure 2C). Cross-channel averaging, as has been done here (Figure 3) and published previously (Curley et al., 2022a) would further increase the signal-to-noise ratio for research or clinical applications.

We note that the hard threshold-based FOOOF was more likely to classify spectra as non-conforming (Z), whereas our CNN classifier had largely preserved accuracy across ABCD categories (Figure 2). Given the CNN classifier was trained directly on human-classified spectra, it may be more capable of capturing the gestalt of borderline spectra and perform accurate classification (against human rating as the gold standard) when a hard-thresholded classifier is not. This likely results in globally lower accuracies for the FOOOF classifier but supports that need for a classifier that can account for nuanced spectral characteristics that are detected by humans to ensure accurate classification.

Several considerations should be noted. The classifier was trained on a relatively small dataset, and the external validation cohort exhibited an imbalanced distribution of ABCD categories, with relatively few *C*-type spectra. This reflects the broader challenge of limited labeled data in this population. While we incorporated synthetic data during training to effectively augment the diversity of spectral patterns (see Supplementary Materials), further validation on larger, more balanced datasets is needed. The classifier was also trained and validated on manually cleaned EEG data. While our application of the classifier to a continuous, raw stretch of EEG demonstrates its robustness in the setting of uncleaned data, the classifier’s performance on data with high levels of artifact has yet to be validated. Increased availability of labeled data is important to better generalize to the full range of EEG phenotypes observed in acute brain injury.

In summary, we developed and evaluated the performance of a learning-based approach for automated ABCD classification, compared it against a state-of-the-art rule-based spectral analysis baseline, and demonstrated its utility for tracking dynamic changes in brain state over time. Our classifier represents a critical step toward translating of the ABCD framework to the bedside and improving clinicians’ ability to diagnose and prognosticate for patients with acute severe brain injuries.

## Data Availability

Data can be requested directly from the authors of respective source studies.

## Abbreviations

CMD: cognitive motor dissociation
CNN: convolutional neural net
EEG: electroencephalography
ICU: intensive care unit
TBI: traumatic brain injury

## Acknowledgements

This work was supported by the Harvard Medical School Office of Scholarly Engagement, NIH National Institute of Neurological Disorders and Stroke (UE5 NS065743, R01 NS138257), the NIH Director’s Office (DP2 HD101400), NIH Shared Instrument Grant S10 RR023043, James S. McDonnell Foundation, and the Chen Institute MGH Research Scholar Award.

## Supplementary Materials

### Synthetic Spectra Generation

Synthetic spectra were generated on-the-fly during training to augment the limited labeled dataset, as illustrated in Figure S1. All spectra were constructed over the 2-24 Hz range and expressed in normalized amplitude units in [0, 1], obtained via min-max scaling within this frequency interval. Each spectrum was initialized with an aperiodic background:

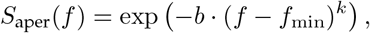

where *k* (exponent) and *b* (offset) were independently and uniformly sampled per instance, with *k* in [0.25, 1] and *b* in [0, 3]. Given a sampled label in {*A, B, C, D, Z*}, we add oscillatory peaks according to label-specific band rules (Figure 1), each modeled as a generalized Gaussian:

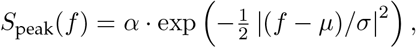

where center frequency *µ*, bandwidth *σ*, and amplitude *α* were independently sampled from band-specific ranges defined in Table S1.

For ABCD categories, peaks were sampled within canonical bands (delta, theta, alpha, beta) consistent with label definitions. For the non-conforming category (*Z*), we simulated structured deviations from ABCD patterns, including paired peaks in (delta, beta), (delta, alpha), and (theta, alpha) bands. Sampling ranges for *µ, σ*, and *α* were derived empirically from the training data. Specifically, all manually annotated peaks were normalized, and the 10^th^ - 90^th^ percentile ranges of their centers, bandwidths, and amplitudes were used to define sampling intervals.

Additive spectral noise was applied as zero-mean Gaussian noise with standard deviations uniformly sampled in [0, 0.02]. Noise was generated at multiple spatial scales (1, 2, 4, and 8 Hz bandwidths) and summed.

**Table S1.** Uniform sampling ranges for oscillatory peak parameters. All amplitudes are normalized to [0, 1].

| Band | Center $\mu$ (Hz) | Width $\sigma$ (Hz) | Amplitude $A$ |
| --- | --- | --- | --- |
| Delta | 1.5 - 3.5 | 0.75 - 2 | 0 - 0.8 |
| Theta | 4 - 8 | 0.75 - 3 | 0.15 - 0.5 |
| Alpha | 8 - 12.5 | 0.75 - 3 | 0.15 - 0.5 |
| Beta | 13 - 21 | 1 - 4 | 0.1 - 0.3 |

### Training Without Synthetic Data

We trained and evaluated the CNN without sampling synthetic data. This resulted in a mean class-balanced recall of 0.72 on the internal cohort and 0.61 on the external co-hort, corresponding to an overall decrease of 0.11 in mean recall relative to the primary results (Table 2).

### FOOOF Hyperparameter Search

In the grid searches to determine key FOOOF hyperparameters, the aperiodic component was modeled using either a fixed or knee formulation. Peak width limits were explored with lower bounds from 1 to 5 Hz (10 evenly spaced values) and upper bounds from 5 to 15 Hz (20 values). Minimum peak height thresholds were sampled between 0 and 1 (50 values), and peak detection thresholds between 0.5 and 4.0 (50 values). The maximum number of peaks was varied from 1 to 6. The optimal configuration used a fixed aperiodic model, peak width limits of 1.3– 10 Hz, a minimum peak height of 0.18, a peak detection threshold of 1.3, and a maximum of 4 peaks.

**Figure S1.**
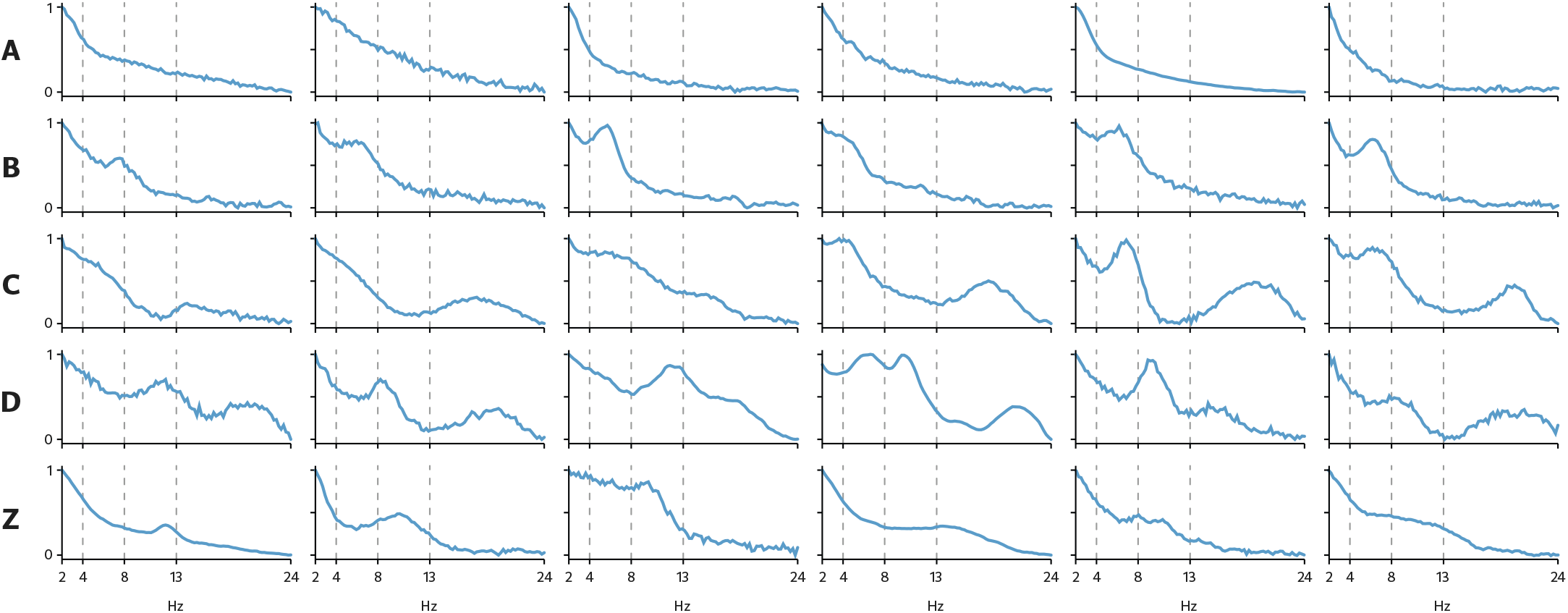
Representative examples of synthesized spectra, with peak structure consistent with the ABCD framework.

## References

Caroline Schnakers, Audrey Vanhaudenhuyse, Joseph Giacino, Manfredi Ventura, Melanie Boly, Steve Majerus, Gustave Moonen, and Steven Laureys. Diagnostic accuracy of the vegetative and minimally conscious state: clinical consensus versus standardized neurobehavioral assessment. BMC neurology, 9 (1):35, 2009.

Nicholas D Schiff. Cognitive motor dissociation following severe brain injuries. JAMA neurology, 72(12):1413–1415, 2015.

Brian L Edlow, Jan Claassen, Nicholas D Schiff, and David M Greer. Recovery from disorders of consciousness: mechanisms, prognosis and emerging therapies. Nature Reviews Neurology, 17(3):135–156, 2021.

Paul M Vespa, W John Boscardin, David A Hovda, David L McArthur, Marc R Nuwer, Neil A Martin, Valeriy Nenov, Thomas C Glenn, Marvin Bergsneider, Daniel F Kelly, et al. Early and persistent impaired percent alpha variability on continuous electroencephalography monitoring as predictive of poor outcome after traumatic brain injury. Journal of neurosurgery, 97(1):84–92, 2002.

Matthew O Hebb, David L McArthur, Jeffry Alger, Maria Etchepare, Thomas C Glenn, Marvin Bergsneider, Neil Martin, and Paul M Vespa. Impaired percent alpha variability on continuous electroencephalography is associated with thalamic injury and predicts poor long-term outcome after human traumatic brain injury. Journal of neurotrauma, 24(4):579–590, 2007.

Peter B Forgacs, Hans-Peter Frey, Angela Velazquez, Stephanie Thompson, Daniel Brodie, Vivek Moitra, Leroy Rabani, Soojin Park, Sachin Agarwal, Maria Cristina Falo, et al. Dynamic regimes of neocortical activity linked to corticothalamic integrity correlate with outcomes in acute anoxic brain injury after cardiac arrest. Annals of clinical and translational neurology, 4(2):119–129, 2017.

Antti Tolonen, Mika OK Särkelä, Riikka SK Takala, Ari Katila, Janek Frantzén, Jussi P Posti, Markus Müller, Mark Van Gils, and Olli Tenovuo. Quantitative eeg parameters for prediction of outcome in severe traumatic brain injury: development study. Clinical EEG and neuroscience, 49(4):248–257, 2018.

Ayham Alkhachroum, Andrey Eliseyev, Caroline A Der-Nigoghossian, Clio Rubinos, Julie Anne Kromm, Elizabeth Mathews, Andrew Bauerschmidt, Kevin Doyle, Angela Velasquez, Jennifer A Egbebike, et al. Eeg to detect early recovery of consciousness in amantadine-treated acute brain injury patients. Journal of Neurology, Neurosurgery & Psychiatry, 91(6): 675–676, 2020.

Peter B Forgacs, Orrin Devinsky, and Nicholas D Schiff. Independent functional outcomes after prolonged coma following cardiac arrest: a mechanistic hypothesis. Annals of neurology, 87(4):618–632, 2020.

Peter B Forgacs, Baxter B Allen, Xian Wu, Linda M Gerber, Srikanth Boddu, Malik Fakhar, Philip E Stieg, Nicholas D Schiff, and Halinder S Mangat. Corticothalamic connectivity in aneurysmal subarachnoid hemorrhage: relationship with disordered consciousness and clinical outcomes. Neurocritical Care, 36(3):760–771, 2022.

Moshgan Amiri, Patrick M Fisher, Federico Raimondo, Annette Sidaros, Melita Cacic Hribljan, Marwan H Othman, Ivan Zibrandtsen, Simon S Albrechtsen, Ove Bergdal, Adam Espe Hansen, et al. Multimodal prediction of residual consciousness in the intensive care unit: the connect-me study. Brain, 146(1):50–64, 2023.

William H Curley, Yelena G Bodien, David W Zhou, Mary M Conte, Andrea S Foulkes, Joseph T Giacino, Jonathan D Victor, Nicholas D Schiff, and Brian L Edlow. Electrophysiological correlates of thalamocortical function in acute severe traumatic brain injury. Cortex, 152:136–152, 2022a.

William H Curley, Angela Comanducci, and Matteo Fecchio. Conventional and investigational approaches leveraging clinical eeg for prognosis in acute disorders of consciousness. In Seminars in Neurology, volume 42, pages 309–324. Thieme Medical Publishers, Inc., 2022b.

David W Zhou, Mary M Conte, William H Curley, Camille A Spencer-Salmon, Camille Chatelle, Eric S Rosenthal, Yelena G Bodien, Jonathan D Victor, Nicholas D Schiff, Emery N Brown, et al. Alpha coherence is a network signature of cognitive recovery from disorders of consciousness. medRxiv, 2024.

Nicholas D Schiff. Recovery of consciousness after brain injury: a mesocircuit hypothesis. Trends in neurosciences, 33(1):1–9, 2010.

Nicholas D Schiff. Mesocircuit mechanisms underlying recovery of consciousness following severe brain injuries: model and predictions. Brain function and responsiveness in disorders of consciousness, pages 195–204, 2016.

Igor Timofeev, F Grenier, M Bazhenov, TJ Sejnowski, and Mircea Steriade. Origin of slow cortical oscillations in deafferented cortical slabs. Cerebral cortex, 10(12):1185–1199, 2000.

Laurie R Silva, Yael Amitai, and Barry W Connors. Intrinsic oscillations of neocortex generated by layer 5 pyramidal neurons. Science, 251(4992):432–435, 1991.

Rodolfo R Llinás, Urs Ribary, Daniel Jeanmonod, Eugene Kronberg, and Partha P Mitra. Thalamocortical dysrhythmia: a neurological and neuropsychiatric syndrome characterized by magnetoencephalography. Proceedings of the National Academy of Sciences, 96(26):15222–15227, 1999.

Rodolfo Llinás, Francisco J Urbano, Elena Leznik, Rey R Ramírez, and Hein JF Van Marle. Rhythmic and dysrhythmic thalamocortical dynamics: Gaba systems and the edge effect. Trends in neurosciences, 28(6):325–333, 2005.

Donald L. Schomer and Fernando H. Lopes da Silva. Niedermeyer’s Electroencephalography: Basic Principles, Clinical Applications, and Related Fields. Oxford University Press, 11 2017. ISBN 9780190228484. doi: 10.1093/med/9780190228484.001.0001.

Michelle J Redinbaugh, Jessica M Phillips, Niranjan A Kambi, Sounak Mohanta, Samantha Andryk, Gaven L Dooley, Mohsen Afrasiabi, Aeyal Raz, and Yuri B Saalmann. Thalamus modulates consciousness via layer-specific control of cortex. Neuron, 106(1):66–75, 2020.

AP Janson, JL Baker, I Sani, KP Purpura, ND Schiff, and CR Butson. Selective activation of central thalamic fiber pathway facilitates behavioral performance in healthy non-human primates. Scientific reports, 11(1):23054, 2021.

Esteban A Fridman, Bradley J Beattie, Allegra Broft, Steven Laureys, and Nicholas D Schiff. Regional cerebral metabolic patterns demonstrate the role of anterior forebrain mesocircuit dysfunction in the severely injured brain. Proceedings of the National Academy of Sciences, 111(17):6473–6478, 2014.

Evan S Lutkenhoff, Jeffrey Chiang, Luaba Tshibanda, Evelyn Kamau, Murielle Kirsch, John D Pickard, Steven Laureys, Adrian M Owen, and Martin M Monti. Thalamic and extrathalamic mechanisms of consciousness after severe brain injury. Annals of neurology, 78(1):68–76, 2015.

David B Fischer, Aaron D Boes, Athena Demertzi, Henry C Evrard, Steven Laureys, Brian L Edlow, Hesheng Liu, Clifford B Saper, Alvaro Pascual-Leone, Michael D Fox, et al. A human brain network derived from coma-causing brainstem lesions. Neurology, 87(23):2427–2434, 2016.

Sean Coulborn, Chris Taylor, Lorina Naci, Adrian M Owen, and Davinia Fernández-Espejo. Disruptions in effective connectivity within and between default mode network and anterior forebrain mesocircuit in prolonged disorders of consciousness. Brain Sciences, 11(6):749, 2021.

Brian L Edlow, Camille Chatelle, Camille A Spencer, Catherine J Chu, Yelena G Bodien, Kathryn L O’Connor, Ronald E Hirschberg, Leigh R Hochberg, Joseph T Giacino, Eric S Rosenthal, et al. Early detection of consciousness in patients with acute severe traumatic brain injury. Brain, 140(9):2399–2414, 2017.

Yelena G Bodien, Judith Allanson, Paolo Cardone, Arthur Bonhomme, Jerina Carmona, Camille Chatelle, Srivas Chennu, Mary Conte, Stanislas Dehaene, Paola Finoia, et al. Cognitive motor dissociation in disorders of consciousness. New England Journal of Medicine, 391(7):598–608, 2024.

Bo Hjorth. An on-line transformation of eeg scalp potentials into orthogonal source derivations. Electroencephalography and clinical neurophysiology, 39(5):526–530, 1975.

GW Thickbroom, FL Mastaglia, WM Carroll, and HD Davies. Source derivation: application to topographic mapping of visual evoked potentials. Electroencephalography and Clinical Neurophysiology/Evoked Potentials Section, 59(4):279–285, 1984.

David J Thomson. Spectrum estimation and harmonic analysis. Proceedings of the IEEE, 70(9):1055–1096, 1982.

Donald B Percival and Andrew T Walden. Spectral analysis for physical applications. cambridge university press, 1993.

Hemant Bokil, Peter Andrews, Jayant E Kulkarni, Samar Mehta, and Partha P Mitra. Chronux: a platform for analyzing neural signals. Journal of neuroscience methods, 192(1):146–151, 2010.

Peter B Forgacs, Mary M Conte, Esteban A Fridman, Henning U Voss, Jonathan D Victor, and Nicholas D Schiff. Preservation of electroencephalographic organization in patients with impaired consciousness and imaging-based evidence of command-following. Annals of neurology, 76(6):869–879, 2014.

William H Curley, Peter B Forgacs, Henning U Voss, Mary M Conte, and Nicholas D Schiff. Characterization of eeg signals revealing covert cognition in the injured brain. Brain, 141(5): 1404–1421, 2018.

Michiel Meys, Glenn van der Lande, Nicolas Lejeune, Olivia Gosseries, Aurore Thibaut, Gang Chen, Mary Conte, Nicholas Schiff, Steven Laureys, Daniele Marinazzo, and Jitka Annen. Eeg dynamic regimes and the contributions of regional glucose uptake in a large cohort of patients with prolonged disorders of consciousness. In 28th Annual Meeting of the Association for the Scientific Study of Consciousness, 2025.

Adam Paszke, Sam Gross, Francisco Massa, Adam Lerer, James Bradbury, Gregory Chanan, Trevor Killeen, Zeming Lin, Natalia Gimelshein, Luca Antiga, et al. Pytorch: An imperative style, high-performance deep learning library. Advances in neural information processing systems, 32, 2019.

Diederik P Kingma and Jimmy Ba. Adam: A method for stochastic optimization. arXiv preprint arXiv:1412.6980, 2014.

Josh Tobin, Rachel Fong, Alex Ray, Jonas Schneider, Wojciech Zaremba, and Pieter Abbeel. Domain randomization for transferring deep neural networks from simulation to the real world. In 2017 IEEE/RSJ international conference on intelligent robots and systems (IROS), pages 23–30. IEEE, 2017.

Jonathan Tremblay, Aayush Prakash, David Acuna, Mark Brophy, Varun Jampani, Cem Anil, Thang To, Eric Cameracci, Shaad Boochoon, and Stan Birchfield. Training deep networks with synthetic data: Bridging the reality gap by domain randomization. In Proceedings of the IEEE conference on computer vision and pattern recognition workshops, pages 969–977, 2018.

Thomas Donoghue, Matar Haller, Erik J Peterson, Paroma Varma, Priyadarshini Sebastian, Richard Gao, Torben Noto, Antonio H Lara, Joni D Wallis, Robert T Knight, et al. Parameterizing neural power spectra into periodic and aperiodic components. Nature neuroscience, 23(12):1655–1665, 2020.

Biyu J He, John M Zempel, Abraham Z Snyder, and Marcus E Raichle. The temporal structures and functional significance of scale-free brain activity. Neuron, 66(3):353–369, 2010.

Adenauer G Casali, Olivia Gosseries, Mario Rosanova, Mélanie Boly, Simone Sarasso, Karina R Casali, Silvia Casarotto, MarieAurélie Bruno, Steven Laureys, Giulio Tononi, et al. A theoretically based index of consciousness independent of sensory processing and behavior. Science translational medicine, 5(198): 198ra105–198ra105, 2013.

Raechelle M Gibson, Davinia Fernández-Espejo, Laura E Gonzalez-Lara, Benjamin Y Kwan, Donald H Lee, Adrian M Owen, and Damian Cruse. Multiple tasks and neuroimaging modalities increase the likelihood of detecting covert awareness in patients with disorders of consciousness. Frontiers in Human Neuroscience, 8:950, 2014.

Andrea Piarulli, Massimo Bergamasco, Aurore Thibaut, Victor Cologan, Olivia Gosseries, and Steven Laureys. Eeg ultradian rhythmicity differences in disorders of consciousness during wakefulness. Journal of neurology, 263(9):1746–1760, 2016.

Sarah Wannez, Lizette Heine, Marie Thonnard, Olivia Gosseries, Steven Laureys, and Coma Science Group Collaborators. The repetition of behavioral assessments in diagnosis of disorders of consciousness. Annals of neurology, 81(6):883–889, 2017.

Joseph T Giacino, Douglas I Katz, Nicholas D Schiff, John Whyte, Eric J Ashman, Stephen Ashwal, Richard Barbano, Flora M Hammond, Steven Laureys, Geoffrey SF Ling, et al. Practice guideline update recommendations summary: disorders of consciousness: report of the guideline development, dissemination, and implementation subcommittee of the american academy of neurology; the american congress of rehabilitation medicine; and the national institute on disability, independent living, and rehabilitation research. Neurology, 91(10): 450–460, 2018.

Athena Demertzi, Enzo Tagliazucchi, Stanislas Dehaene, Gustavo Deco, Pablo Barttfeld, Federico Raimondo, Charlotte Martial, Davinia Fernández-Espejo, Benjamin Rohaut, HU Voss, et al. Human consciousness is supported by dynamic complex patterns of brain signal coordination. Science advances, 5(2):eaat7603, 2019.

Daniel Kondziella, Andreas Bender, Karin Diserens, Willemijn van Erp, Anna Estraneo, Rita Formisano, Steven Laureys, Lionel Naccache, S Ozturk, Benjamin Rohaut, et al. European academy of neurology guideline on the diagnosis of coma and other disorders of consciousness. European journal of neurology, 27(5):741–756, 2020.

Jan Claassen, Kevin Doyle, Adu Matory, Caroline Couch, Kelly M Burger, Angela Velazquez, Joshua U Okonkwo, Jean-Rémi King, Soojin Park, Sachin Agarwal, et al. Detection of brain activation in unresponsive patients with acute brain injury. New England Journal of Medicine, 380(26):2497–2505, 2019.

